# Does the use of intraoperative CT scan improve outcomes in Orthopaedic surgery? A protocol for systematic review and meta-analysis

**DOI:** 10.1101/2021.03.01.21252670

**Authors:** Vishal Kumar, Vishnu Baburaj, Sandeep Patel, Siddhartha Sharma, Raju Vaishya

## Abstract

**Background:** Intraoperative 3D imaging is a valuable tool to visualise complex anatomical structures and position implants optimally. This is an emerging technology and there is a paucity in literature of articles describing the same.

**Objectives:** This systematic review aims to compare the outcome of surgeries done under 3D imaging to those done under 2D fluoroscopy by analysing the evidence in literature.

**Methods:** A systematic review will be conducted adhering to the PRISMA guidelines. Electronic databases of PubMed, Embase and Scopus will be searched with a pre-defined search string. A manual search will also be conducted on the references of the included articles. Original research articles published in English that directly compare the outcomes of intraoperative 3D imaging with fluoroscopy will be included. Review articles, conference abstracts and animal studies will be excluded. Outcome measures will be extracted from the selected studies and analysed with the help of appropriate software.

## 1. Background

Intraoperative radiography is commonly used in Orthopaedic surgery. Historically, 2D fluoroscopy has been extensively used for this purpose. 2D imaging, however, falls short when visualisation of complex structures are required, such as spine, pelvis etc that have complex anatomy and extensive soft tissue coverage. As a solution for this, intraoperative 3D imaging (iCT) was introduced, as early as in 2001 (1). This maybe achieved by a motorised C-Arm which rotates automatically about the area of interest, capturing a large number of images which are then compiled in 3D format by a software and made available to the operating team. More recently, mobile CT scanners have been developed that are very similar to conventional CT scanners, and can be used intraoperatively for better quality 3D imaging.

## 2. Need for review

There is limited literature on the use of iCT in Orthopaedics and there are no systematic reviews done till date. Hence this systematic review aims to give a comprehensive overview of intraoperative 3D imaging and compare it with conventional 2D fluoroscopy.

## 3. Objectives

- To provide a complete outline on the current applications of iCT
- To compare outcomes of surgeries done under iCT with those done under conventional fluoroscopy

## 4. PICO framework for the study

a. Participants : Human subjects
b. Intervention : Surgeries in which iCT was employed
c. Control : Surgeries done under conventional 2D fluoroscopy
d. Outcome : Accuracy of implant positioning and reduction, mean surgical time, mean blood loss

## 5. Methods

This systematic review and meta-analysis will be conducted adhering to the Preferred Reporting Items for Systematic Reviews and Meta-analysis (PRISMA) guidelines.

a. Review Protocol A protocol of the review will be prepared according to the PRISMA-P guidelines.
b. Eligibility Criteria Any original research on human subjects that compare iCT with fluoroscopy will be included. Studies which have described the use of iCT with appropriate data on outcomes will also be included. Studies in languages other than English, animal studies, review articles, conference abstracts will be excluded.
c. Information Sources & Literature search A literature search will be conducted on the following electronic databases of: PubMed, Embase and Scopus using the search string “Intraoperative AND CT OR (computed tomography) AND Orthopaedics”. A manual reference search of the included articles will also be carried out for potentially eligible articles. Articles published from inception till date of search will be included.
d. Study Selection Two independent authors will screen the title and abstract of all articles obtained from the initial search. Full text of potentially eligible articles will be obtained and evaluated according to the inclusion and exclusion criteria. Any conflict of judgement with regard to inclusion or exclusion of any articles will be resolved by discussion between the authors.
e. Data Collection & Data Items Date will be extracted on specially designed data collection excel spreadsheets, and will be cross checked for accuracy. The data that will be collected will include:
  - Name of first author and year of publication
  - Study design
  - Number of participants
  - Average duration of surgery
  - Mean blood loss
  - Number of accurate implants positioned to total number of implants
  - Outcome at follow up
  - Conclusions of authors
f. Outcome Measures The following outcome measures will be evaluated:
  - Accuracy of implant placement
  - Mean surgical duration
  - Average blood loss
  - Rate of revision surgery
g. Data Analysis and Synthesis Qualitative and qualitative synthesis will be carried out. Quantitative analysis will be done to ascertain the pooled estimates of outcome parameters if it is reported by more than 3 studies. Meta-analysis will be carried out using RevMan version 5.4 with random-effects model with 95% confidence intervals. Results will be depicted with the help of forest plots.
h. Risk of bias Risk of bias assessment will be done using MINORS tool for observational studies and ROB 2.O tool for randomised control trials (2, 3).

## Data Availability

All data referred to in the manuscript will be available as supplementary files along with the final article.

## References

1. Keil H, Beisemann N, Schnetzke M, Vetter SY, Grützner PA, Franke J. First experiences with the Airo mobile intraoperative CT scanner in acetabular surgery-An analysis of 10 cases. Int J Med Robot. 2019 Apr;15(2):e1986.

2. Slim K, Nini E, Forestier D, Kwiatkowski F, Panis Y, Chipponi J. Methodological index for non□randomized studies (MINORS): development and validation of a new instrument. ANZ journal of surgery. 2003 Sep;73(9):712–6.

3. Sterne JAC, Savović J, Page MJ, Elbers RG, Blencowe NS, Boutron I, Cates CJ, Cheng H-Y, Corbett MS, Eldridge SM, Hernán MA, Hopewell S, Hróbjartsson A, Junqueira DR, Jüni P, Kirkham JJ, Lasserson T, Li T, McAleenan A, Reeves BC, Shepperd S, Shrier I, Stewart LA, Tilling K, White IR, Whiting PF, Higgins JPT. RoB 2: a revised tool for assessing risk of bias in randomised trials. BMJ 2019; 366: 4898.

